# Cost-Benefit Analysis of a Distracted Pedestrian Intervention

**DOI:** 10.1101/2022.08.14.22278757

**Authors:** Md Jillur Rahim, David C. Schwebel, Ragib Hasan, Russell Griffin, Bisakha Sen

## Abstract

**Objective:** Cellphone ubiquity has increased distracted pedestrian behavior and contributed to growing pedestrian injury rates. A major barrier to large-scale implementation of prevention programs is unavailable information on potential net monetary benefits. We evaluated net economic benefits of StreetBit, a program that reduces distracted pedestrian behavior by sending warnings from intersection-installed Bluetooth beacons to distracted pedestrians’ smartphones.

**Methods:** Three data sources were used: (1) fatal, severe, non-severe pedestrian injury rates from Alabama’s electronic crash-reporting-system; (2) expected costs per fatal, severe, non-severe pedestrian injury – including medical cost, value of statistical life, work-loss cost, quality-of-life cost – from CDC; and (3) prevalence of distracted walking from extant literature. We computed and compared estimated monetary costs of distracted walking in Alabama and monetary benefits from implementing StreetBit to reduce pedestrian injuries at intersections.

**Results:** Over 2019-2021, Alabama recorded an annual average of 31 fatal, 83 severe, and 115 non-severe pedestrian injuries in intersections. Expected costs/injury were $11 million, $339,535, and $93,877, respectively. The estimated range of distracted walking prevalence is 25%-40%, and StreetBit demonstrates 19.1% (95%CI: 1.6%-36.0%) reduction. These figures demonstrate potential annual cost savings from using interventions like StreetBit statewide ranging from $18.1-$29 million. Potential costs range from $3,208,600 (beacons at every-fourth urban intersection) to $6,359,200 (every other intersection).

**Conclusions:** Even under the most parsimonious scenario (25% distracted pedestrians; densest beacon placement), StreetBit yields $11.8 million estimated net annual benefit. Existing data sources can be leveraged to predict net monetary benefits of distracted pedestrian interventions like StreetBit and facilitate large-scale intervention adoption.

**SUMMARY:** *What is already known in this topic:* Smartphone-related distraction is a likely contributing factor to the increasing rate of pedestrian fatalities and injuries in the US. However, interventions to reduce pedestrian distraction have not been widely adapted.

*What this study adds:* One barrier to widespread adaption is lack of information on benefits versus costs. This study examines the economic costs and benefits of an intervention that reduces distracted walking to increase pedestrian safety, and provides a template showing how existing data sources can be leveraged to do similar analyses for other interventions designed to reduce pedestrian safety.

*How this study might affect research, practice or policy:* The template developed in this study can facilitate large-scale implementation of any intervention designed to prevent pedestrian fatalities and injuries by providing policymakers information on net benefits of the intervention.

## INTRODUCTION

Over 6,500 Americans died in a pedestrian crash in 2020, according to the most recent data available from the National Center for Statistics and Analysis (NCSA, 2022). This represents a 47% increase from pedestrian fatalities reported in 2011 (NCSA, 2014), in contrast to the 20% increase in total traffic fatalities over the same period. This dramatic increase in pedestrian injury deaths, which remains present after adjusting for population changes, is attributed to various causes. One likely contributing factor is the increasing use of smartphones by pedestrians in and near traffic (Fischer, 2015; Ralph & Girardeau, 2020; Retting & Rothenberg, 2015).

Cognitive-perceptual research repeatedly demonstrates that smartphone use negatively impacts pedestrian safety (Simmons et al., 2020; Stavrinos et al., 2011). Experts cite three components of distraction: (a) visual inattention, which results from the pedestrian’s visual attention being diverted to smartphone screens instead of the surrounding traffic environment; (b) auditory inattention, which results from the pedestrian’s auditory attention being diverted to smartphone music or conversations instead of the surrounding traffic environment; and (c) cognitive inattention, which results from the pedestrian’s cognitive attention being diverted to the smartphone and its contents rather than the cognitively complex surrounding traffic environment. One recent meta-analysis offers respective effect sizes of *r* = .17 (95% CI .12, .22; Cohen’s d = .34), *r* = .34 (95% CI .23, .46; Cohen’s d = .73), and *r* = .18 (95% CI -.12, .49; Cohen’s d = .37), demonstrating the impact of talking, texting/browsing, and music listening on hits or close calls in simulated pedestrian crossings (Simmons et al., 2020).

Despite evidence that distraction reduces pedestrian safety and that the sharp increase in pedestrian fatalities in the United States over the past decade is attributed partly to distracting behavior by pedestrians, there have been comparatively few attempts to develop and evaluate effective and cost-efficient strategies to reduce distracted pedestrian behavior. Efforts to place warning signs or lights on sidewalks or in crossing areas show initial promise of effectiveness in some trials (Larue & Watling, 2021) but mixed results in other attempts (Barin et al., 2018; Kim et al., 2021; Violano et al., 2015). “Distracted walking laws” have been implemented in just a few jurisdictions around the world, and public health initiatives to reduce distracted walking have not been rigorously evaluated (Schwebel, McClure, & Porter, 2017).

One barrier to large-scale implementation by municipalities, cities, or states of interventions that show promise in pilot or experimental studies is uncertainty around whether the benefits, when translated to monetary terms, will justify the costs. This creates the proverbial “chicken and egg” problem, whereby large-scale implementation does not occur due to a lack of information on monetary costs versus benefits, and because large-scale implementations do not happen, in turn, no data is generated that will permit evaluation of costs and benefits. What may help break this impasse is a template that enables researchers to estimate monetary costs and benefits primarily using existing data. This manuscript provides such a template – using a recently-described strategy, StreetBit, -- as an example of an intervention that shows promise of reducing distracted pedestrian behavior. Drawing from preliminary empirical findings on the effectiveness of StreetBit in reducing distracted pedestrian behavior and combining that with existing data on the incidence and costs of pedestrian injury, this analysis shows whether StreetBit might be a cost-efficient strategy to reduce the monetary fallout from distracted pedestrian behavior.

StreetBit directly warns distracted pedestrians on their smartphones as they approach a street corner while distracted (Hasan et al., 2021; Schwebel et al., 2021). Bluetooth beacons are installed on street corners and send unidirectional signals to pedestrians using their smartphones when they come in contact with the beacons. Pedestrians looking at their phones receive a visual warning, and those listening to their phones receive an auditory message.

Preliminary testing of StreetBit was promising (Schwebel et al., 2021). Before large-scale dissemination of programs like StreetBit, however, a framework for empirically estimating the costs and benefits of implementing the program that gives local leaders and policymakers information on the return on investment of the large-scale adoption to secure their cooperation.

To accomplish our goal, we gathered data estimating the cost of installing StreetBit at intersections across the state of Alabama and the costs of installing the software on pedestrians’ phones. Together, these data represent the costs of the program. We also estimated the economic benefits of the program based on the reduction in costs of pedestrian injuries prevented. The aim was to build a cost-benefit analysis framework whereby the benefits of distracted pedestrian prevention programs like StreetBit can be estimated by leveraging and synthesizing existing data from appropriate sources. Beyond informing stakeholders and policymakers about the benefits versus costs of large-scale adaption of StreetBit, this framework has the advantage in that it can be used and adapted by other researchers considering distracted pedestrian interventions and offers guidance on predicting the economic benefit of large-scale dissemination of those interventions.

## METHODS

### Study Location

We focused our analysis on Alabama; the US state ranked as the second most dangerous state for walkers on the streets by Smart Growth America and the National Complete Streets Coalition (Smart Growth America, 2021).

### Overview of Methods

We estimated costs and benefits to accomplishing our goal of estimating the benefit of StreetBit in terms of monetary savings. To estimate costs, we built a logic model that outlined the key components of costs associated with pedestrian fatality and injury and the baseline prevalence of such fatality and injury (See Figure 1). To populate the costs of pedestrian injury and fatality in the conceptual model, we extracted and utilized data from two sources: (a) eCrash, an electronic traffic crash reporting system for the state of Alabama, which was used to obtain counts of pedestrian injury in the state, including fatal injuries and injuries by the level of severity, and (b) the CDC Web-based Injury Statistics Query and Reporting System (WISQARS) Cost of Injury Reports, which was used to obtain costs associated with pedestrian fatal injuries and injuries of different level of severity. Each dataset was restricted to pedestrian injuries and fatalities occurring in the intersections in the state of Alabama.

Next, we considered published reports on the proportion of pedestrians distracted by mobile devices while walking (Wells et al., 2018) and on the effectiveness of StreetBit in reducing such distractions (Schwebel et al., 2021). These data allowed us to estimate the potential number of fatalities and injuries that would be prevented from the large-scale implementation of StreetBit across Alabama. The monetary savings from these prevented fatalities/injuries constitute the predicted benefit of StreetBit.

### Data Sources

#### Fatal & Non-fatal Pedestrian Injuries

Fatal and nonfatal pedestrian injury data in Alabama were collected from the state’s electronic crash reporting system, developed by the University of Alabama Center for Advanced Public Safety. We conservatively restricted our analysis to injuries occurring at intersections only, as it is unknown what effect programs like StreetBit might have on injuries occurring at non-intersection locations. We used an average of the three most recently-released years of data (2019, 2020, and 2021) for our analysis.

Nonfatal injuries were categorized into two types, severe and non-severe. Severe injuries included those that the eCrash system categorized as “serious” injuries, and minor injuries were classified as non-severe injuries.

#### Costs of Injury

For all three categories of injuries (fatal, severe nonfatal, non-severe nonfatal), costs of the injury were obtained from CDC WISQARS (CDC, 2022). CDC WISQARS provides mean national costs for fatal pedestrian injuries, severe injuries (defined as injuries requiring hospitalization), and non-severe injuries (defined as injuries where a patient was treated and released at the emergency department). Estimated costs for fatalities include costs for medical care and the statistical value of life, and costs for nonfatalities include medical care costs, work loss costs, and quality of life costs. Because the latest data available were from 2020, we inflation-adjusted medical costs to 2021 using the Consumer Price Index (CPI) for medical care, and inflation-adjusted value of statistical life, work loss costs, and quality of life cost using the general CPI. WISQARS data were used because corresponding cost data specific to Alabama are unavailable in the eCrash system.

#### Percentage of Distracted Pedestrians

No rigorous scientific evidence has been published concerning what percent of pedestrians experiencing an injury were distracted, probably because many injuries occur without reliable witnesses to document whether the pedestrian was distracted or not. Thus, we assumed that the percent of pedestrians experiencing an injury because they were distracted would reflect the overall percent of pedestrians observed to be distracted. In reality, distracted pedestrians may be more likely to be injured than undistracted pedestrians, so our estimates are conservative.

A recent observational study conducted at the University of Alabama at Birmingham and Old Dominion University found that 41.2% of pedestrians were distracted by handheld mobile devices (Wells et al., 2018). However, that study was conducted on urban college campuses where distraction rates may be higher than in other settings. Recently-collected data from our Alabama laboratory found a distraction rate of 30.4% among pedestrians across multiple locations, including entertainment districts, a downtown business district, and near middle and high schools and a university campus (Schwebel et al., 2022). To accommodate the uncertainty inherent in these figures, we calculated the costs of distracted walking under the assumption of 25%, 30%, 35%, or 40% of injured pedestrians being distracted.

#### Total Costs of Distracted Walking

The <u>total cost</u> of distracted walking is represented by *C*_*Q*_, calculated as follows:

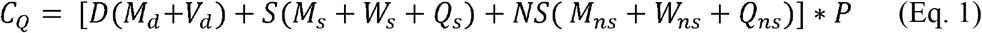

where D represents the total number of pedestrian deaths, S represents the total number of severe pedestrian injuries, and NS represents the total number of non-severe pedestrian injuries; *M*_*d*_, *M*_*s*_, and *M*_*ns*_, respectively represent medical costs associated with pedestrian deaths, severe injuries, and non-severe injuries; *V*_*d*_ represents the value of statistical life for pedestrian deaths; *W*_*s*_, *and W*_*ns*_, respectively represent work loss cost associated with severe injuries and non-severe pedestrian injuries; *Q*_*s*_ & *Q*_*ns*_, represents quality-of-life costs for severe and non-severe nonfatal injuries; and *P* denotes the predicted proportion of pedestrians who were distracted when experiencing an injury.

#### Benefit of StreetBit

The monetary benefit of StreetBit is defined as the cost savings from the expected reductions in pedestrian injury. We used results from Schwebel, Hasan et al. (2021) and the standard mathematical formula to derive the “marginal effect” from odds ratios in logit models (Norton & Dowd, 2018) to obtain an estimated percentage of reduction in distracted walking among pedestrians who were deemed to be most distracted at baseline. This calculation used the assumption that concurrent percentage reductions in pedestrian injuries would be distributed proportionately across fatal, severe nonfatal, and non-severe nonfatal injuries and corresponding decreases in the associated costs.

#### Costs of StreetBit

To estimate the costs of StreetBit, we made the following assumptions. First, we calculated the physical costs of beacons at $15/beacon and their current market price and assumed, based on existing research, that an average of 10 beacons would have to be placed at each street corner (Hasan et al., 2021). The operational lifetime of one StreetBit beacon is four years. However, we assumed 10% of beacons would need replacement annually due to theft or vandalism. Further, the batteries need replacing yearly, at the cost of $1/battery. We also assumed that one employee would dedicate 50 percent of their annual time toward maintaining the beacons and assumed that, inclusive of benefits, in Alabama would cost $40,000. In addition, costs for cloud servers to store data would be $1,500 per month or $18,000 per year.

Given basic psychological principles that a variable ratio learning schedule creates effective learning, behavior change, and resistance to extinction (Baron, 2001; Herrnstein & Heyman, 1979), we assumed StreetBit would not need to be placed at every single intersection to achieve the desired outcome of stopping distracted pedestrian behavior. Without existing empirical evidence to guide us, we computed three scenarios: placing StreetBit beacons at every second, every third, or every fourth intersection in all urban locations across Alabama. We restricted placement to urban locations based on the assumption that they have greater population density and higher frequency of pedestrian activities, and extant evidence that pedestrian injuries involving electronic devices such as headphones overwhelmingly occur in urban counties compared to rural counties (Lichenstein et al., 2012).

#### Patient and Public Involvement

Patients or the public were not involved in the design, conduct, reporting, and dissemination plans of this research. We followed the Consolidated Health Economic Evaluation Reporting Standards 2022 (CHEERS 2022) checklist for reporting the study.

## RESULTS

There were 115, 99, and 126 fatal pedestrian injuries in Alabama in 2019, 2020, and 2021 respectively, averaging 113 deaths annually in the past three years. Of these fatalities, 19, 38, and 36 occurred in the intersections in 2019, 2020, and 2021 respectively (average = 31 per year). The state recorded 92, 73, and 84 severe nonfatal pedestrian injuries in intersections in 2019, 2020, and 2021 respectively, averaging 83 severe injuries annually. Finally, 130, 108, and 107 non-severe nonfatal pedestrian injuries occurred in the intersections in 2019, 2020, and 2021, respectively, averaging 115 non-severe pedestrian injuries in Alabama annually.

The average medical costs and value of statistical life associated with each fatal pedestrian injury in 2020 dollars were $14,169 and $10.46 million, respectively, and $14,311 and $10.98 million after adjusting for inflation. The average medical cost, work loss cost, and quality of life cost were $99,647, $22,406, and $205,470 for severe non-fatal pedestrian injuries and $9,184, $2,213, and $78,360 for non-severe non-fatal injuries. Adjusting these for 2021 figures using CPI medical care, the 2021 medical costs per severe non-fatal injury and non-severe non-fatal injury were $100,643 and $9,276, respectively. Adjusting for 2021 using general CPI, work loss costs associated with severe non-fatal injury and non-severe non-fatal injury were $23,148 and $2,324, and quality of life costs were $215,744 and $82,278, respectively.

Based on previous findings (Schwebel et al., 2022; Wells et al., 2018) regarding the proportion of distracted pedestrians, we permitted *P* in Eq 1 to vary between 25% and 40%. Based on results from Schwebel, Hasan et al. (2021), we assumed the StreetBit program would reduce distracted pedestrian behavior by an average of 19.1% (95% CI: 1.6%-36.0%) across all categories of pedestrian injuries (fatal, nonfatal severe, nonfatal non-severe).

As shown in Table 1, with the most conservative estimate of 25% of injured pedestrians being distracted, the potential annual savings from implementing StreetBit statewide in Alabama are $18,139,937, with a 95% CI ranging from $1,519,576 to $34,190,457. If the less conservative estimate of 40% of injured pedestrians being distracted is used, the estimate increases to $29,023,899 (95% CI: $2,431,321 to $54,704,732).

**Table 1:** Costs of Distracted Walking & Potential Savings Due to StreetBit in Alabama.

|  | Percentage of Distracted Pedestrians |  |  |  |
| --- | --- | --- | --- | --- |
|  | 25% | 30% | 35% | 40% |
| Fatalities | \$85,229,158 | \$102,274,989 | \$119,320,821 | \$136,366,653 |
| Severe Injury | \$7,045,357 | \$8,454,428 | \$9,863,500 | \$11,272,571 |
| Non-severe Injury | \$2,698,978 | \$3,238,773 | \$3,778,569 | \$4,318,365 |
| Cost of Distracted Pedestrian Injuries | \$94,973,493 | \$113,968,191 | \$132,962,890 | \$151,957,588 |
| Decrease in Distractions using StreetBit (95% CI) | 19.1%<br>(1.6% - 36%) |  |  |  |
| Potential Savings due to StreetBit (19.1% of Total Cost of Distracted Pedestrian Injuries) | \$18,139,937 | \$21,767,924 | \$25,395,912 | \$29,023,899 |
| Potential Savings (95% C.I. Lower Limit) | \$1,519,576 | \$1,823,491 | \$2,127,406 | \$2,431,321 |
| Potential Savings (95% C.I. Upper Limit) | \$34,190,457 | \$41,028,549 | \$47,866,640 | \$54,704,732 |

Note: 95% C.I. lower limit is 1.6% of the Total Cost of Distracted Pedestrian Injuries occurring in the intersections, and the upper limit is 36% of the Total Cost of Distracted Pedestrian Injuries. A detailed breakdown of costs can be found in Table A1.

Alabama has an estimated 168,031 intersections across 461 urban areas, which include cities, large towns, and small towns (Boeing, 2020). Table 2 represents a cost breakdown for installing Bluetooth Beacons at an individual intersection. Based on the above estimates, placing beacons in one of every four urban intersections in Alabama would cost $3,208,600 annually, including the fixed costs. Comparable figures to place them in one of every three or one of every two intersections are $4,258,825 and $6,359,200, respectively.

**Table 2:** Bluetooth Beacons Installation Cost Breakdown Per Intersection.

|  | **Lifetime Cost**<br>**(4-year)** | **Annual Cost** |
| --- | --- | --- |
| 10 beacons at \$20/beacon | \$200 | \$50 |
| 1 beacon replaced each year (4 beacons replaced in 4 years) | \$80 | \$20 |
| Batteries replaced annually | \$12 | \$3 |
| **Total Cost Per Intersection** | **\$300** | **\$75** |

Note: The half-time annual employee salary is a fixed cost of the program. It is not included in the table of per beacon costs but is included in the final cost-benefit analyses.

Even with our most conservative estimate of 25% distracted pedestrians, placing a beacon in every fourth intersection would lead to an annual net benefit of $14.9 million for the state of Alabama. Placing beacons in every second or every third intersection would result in a net savings of $13.8 million and $11.78 million, respectively. If the higher estimate of 40% distracted pedestrians is used, then placing beacons at every second, third, or fourth intersection yields net benefits of $22.7 million, $24.8 million, and $25.8 million, respectively. Net benefits under different scenarios are shown in the Appendix.

## DISCUSSION

A rich literature confirms the benefits of policies designed to prevent distracted driving – such as bans on texting while driving – in terms of preventing fatalities and injuries (Ferdinand et al., 2019; Ferdinand et al., 2015). Such information, in turn, permits calculating the costs and effectiveness of interventions designed to prevent motor vehicle injuries (Ecola et al., 2018). However, despite growing evidence of distracted pedestrian behavior due to ubiquitous cellphone use, and its causal link to pedestrian injury, there is a lack of information and tools to compute the costs and effectiveness of interventions to reduce distracted pedestrian behavior. This poses a significant barrier to adopting such interventions by municipalities or states. Our paper offers a roadmap for how existing data and scientific findings can be leveraged to predict the net monetary benefits – i.e., benefits less the program costs – of such interventions by implementing the StreetBit program across Alabama as an example.

Our analysis of the net monetary benefits of implementing a program like StreetBit to reduce distracted pedestrian behavior provides strong evidence that the program is financially beneficial for states or local municipalities to install. Even in the most conservative of estimated scenarios, the combined financial benefit of reduced medical and work loss costs outweighs the costs of installing and maintaining the program.

Programs like StreetBit offer a compelling behavior change strategy because they disrupt a pedestrian’s typical behavior at the moment they are engaging in a risk (Hasan et al., 2021; Schwebel et al., 2021). As a pedestrian approaches an intersection while distracted, StreetBit provides a direct and clear reminder to attend to traffic while crossing the street rather than allowing oneself to be distracted by a smartphone. Similar to injury prevention programs proven to be effective, like smoke detectors and emergency exit signs, the intervention is largely passive; it occurs in the background and provides a reminder to the individual at the moment of risk, encouraging safe behavior. It can arguably be considered “intrusive,” but similar interventions, which are designed to prevent dangerous behavior at the time and location of risk, are successful in other domains, such as the issuance of seat belt reminders in automobiles and the construction of fences around backyard swimming pools (Gielen, Sleet, & DiClemente, 2006; Krafft et al., 2006; Lie et al., 2008).

Our cost savings analysis was purposely conservative and likely underestimates the total costs associated with distracted walking for at least four reasons. First, we restricted our analysis to injuries occurring in the intersections only. We also excluded “possible injuries” occurring in the intersections for which severity could not be determined. There were 86, 49, and 68 “possible injuries” occurring in the intersections in 2019, 2020, and 2021 respectively, averaging 68 possible injuries in the past three years. Second, our calculations omit costs for police and emergency personnel who attend to crash sites and victims, administrative costs of filing records following pedestrian-vehicle crashes, and possible disruptions and delays for other pedestrians and vehicles around the location where crashes occur. Third, our calculations omit the more intangible costs of pedestrian injuries, such as emotional trauma or shock. Fourth, the count of pedestrian injuries may be an undercount of collisions involving a pedestrian where injuries were perceived as non-severe or did not have a crash report filed by a law enforcement agency. Fifth, we assumed the rate of injured pedestrians who were distracted would be proportionate to the total number of pedestrians who are observed to be distracted, whereas it seems likely that injured pedestrians are more likely to be distracted than the overall number of pedestrians. Thus, the cost savings and net benefits from the wide adoption of a program like StreetBit may be higher than what we have estimated here.

Our analysis had some limitations. First, we were bound by the data available and therefore made various estimates and assumptions in our calculations. For example, we used national averages for costs associated with fatal, severe, and non-severe injuries since there were no available corresponding figures specific to Alabama. Relatedly, publicly available nonfatal pedestrian injury estimates such as those in WISQARS are based on a probability sample that did not allow state-specific estimates. In all instances when there was a choice, we erred toward the most conservative estimates; hence, the actual net monetary benefits may be higher than those we report. Second, our estimate of the impact of StreetBit on distracted walking was based on research conducted at a large urban university. College students are especially prone to be distracted by their phones (Stavrinos et al., 2011), and younger pedestrians are less cautious when crossing streets than older pedestrians (Aghabayk et al., 2021). Larger clinical trials will be needed to test the impact of StreetBit on distracted walking in other settings and among other general populations. Third, we assumed costs of implementing StreetBit and medical and work loss costs would be stable over time. Inflation is likely to impact all costs similarly. However, the costs of technology (e.g., beacons and batteries) may decrease or be resistant to inflationary trends, whereas medical and work loss costs may increase more rapidly. Finally, we did not include the costs of promoting and placing StreetBit or a similar program on pedestrians’ smartphones.

In conclusion, we found that StreetBit is cost-effective for local municipalities to improve safety. Equally importantly, this study provided a template that can be used by other researchers and stakeholders to evaluate the cost-effectiveness of large-scale implementation of pilot interventions that reduce distracted walking.

## Data Availability

All data used in this manuscript are publicly available. the sources are described in the manuscript.

## Acknowledgments

Research reported in this publication was supported by the National Science Foundation under Grant Award Number 1952090 and the Eunice Kennedy Shriver National Institute of Child Health & Human Development of the National Institutes of Health under Award Number R21HD095270. The content and any opinions, findings, and conclusions or recommendations expressed in this material are solely the responsibility of the authors and do not necessarily represent the official views of the National Science Foundation or the National Institutes of Health. The authors have no competing interests to declare. Communication regarding this article can be directed to

## Notes

### Competing Interest Statement

The authors have declared no competing interest.

