## Appendix for "Cost-Benefit Analysis of a Distracted Pedestrian Intervention"

**Table A1: Costs of Distracted Walking & Potential Savings Due to StreetBit in Alabama**

|  |  | **Percentage of Distracted Pedestrians** | | | |
| --- | --- | --- | --- | --- | --- |
|  | **Costs** | **25%** | **30%** | **35%** | **40%** |
| Death | Medical Costs | $110,908 | $133,089 | $155,271 | $177,453 |
|  | Value of Statistical Life | $85,118,250 | $102,141,900 | $119,165,550 | $136,189,200 |
| Severe Injury | Medical Costs | $2,088,352 | $2,506,022 | $2,923,693 | $3,341,363 |
|  | Work Loss Costs | $480,327 | $576,393 | $672,458 | $768,524 |
|  | Quality of Life Costs | $4,476,678 | $5,372,013 | $6,267,349 | $7,162,684 |
| Non-severe Injury | Medical Costs | $266,680 | $320,016 | $373,353 | $426,689 |
|  | Work Loss Costs | $66,805 | $80,166 | $93,527 | $106,888 |
|  | Quality of Life Costs | $2,365,493 | $2,838,591 | $3,311,690 | $3,784,788 |
| Total Cost of Distracted Pedestrian Injuries | | $94,973,493 | $113,968,191 | $132,962,890 | $151,957,588 |
| Decrease in Distractions using StreetBit  (95% CI) | | 19.1%  (1.6% - 36%) | 19.1%  (1.6% - 36%) | 19.1%  (1.6% - 36%) | 19.1%  (1.6% - 36%) |
| Potential Savings due to StreetBit (19.1% of Total Cost of Distracted Pedestrian Injuries) | | $18,139,937 | $21,767,924 | $25,395,912 | $29,023,899 |
| Potential Savings (95% C.I. Lower Limit) | | $1,519,576 | $1,823,491 | $2,127,406 | $2,431,321 |
| Potential Savings (95% C.I. Upper Limit) | | $34,190,457 | $41,028,549 | $47,866,640 | $54,704,732 |

**Table A2: Total Cost of Bluetooth Installation**

| Installation Scenarios | Number of Intersections | Beacon Cost Per Intersection | Cost of Beacon | Fixed Cost | Total Cost |
| --- | --- | --- | --- | --- | --- |
| One in Four Intersections | 42008 | $75 | $3,150,600 | $58,000 | $3,208,600 |
| One in Three Intersections | 56011 | $75 | $4,200,825 | $58,000 | $4,258,825 |
| One in Two Intersections | 84016 | $75 | $6,301,200 | $58,000 | $6,359,200 |

**Table A3: Net Savings if Beacons are Installed in One in Four Intersections**

| % of Distracted Pedestrians | Potential Savings | Cost of Beacons | Net Savings |
| --- | --- | --- | --- |
| 25% | $18,139,937 | $3,208,600 | $14,931,337 |
| 30% | $21,767,924 | $3,208,600 | $18,559,324 |
| 35% | $25,395,912 | $3,208,600 | $22,187,312 |
| 40% | $29,0238,99 | $3,208,600 | $25,815,299 |

**Table A4: Net Savings if Beacons are Installed in One in Three Intersections**

| % of Distracted Pedestrians | Potential Savings | Cost of Beacons | Net Savings |
| --- | --- | --- | --- |
| 25% | $18,139,937 | $4,258,825 | $13,881,112 |
| 30% | $21,767,924 | $4,258,825 | $17,509,099 |
| 35% | $25,395,912 | $4,258,825 | $21,137,087 |
| 40% | $29,0238,99 | $4,258,825 | $24,765,074 |

**Table A5: Net Savings if Beacons are Installed in One in Two Intersections**

| % of Distracted Pedestrians | Potential Savings | Cost of Beacons | Net Savings |
| --- | --- | --- | --- |
| 25% | $18,139,937 | $6,359,200 | $11,780,737 |
| 30% | $21,767,924 | $6,359,200 | $15,408,724 |
| 35% | $25,395,912 | $6,359,200 | $19,036,712 |
| 40% | $29,0238,99 | $6,359,200 | $22,664,699 |
